# Influenza hospital admissions prevented by vaccination: a transmission dynamic analysis of the 2022/2023 and 2023/2024 programmes in England

**DOI:** 10.1101/2025.05.11.25327378

**Authors:** Edwin van Leeuwen, Neil Wilkins, Conall H. Watson

## Abstract

We analyse the impact of the seasonal influenza vaccination programme over the 2022/2023 and 2023/2024 winter seasons in England, the first two full seasons since the re-emergence of influenza following the COVID-19 pandemic. Our approach adapts an existing age and risk-stratified SEEIIR dynamic transmission model, by additionally fitting to hospital admission surveillance data and allowing for changes in mixing patterns during school holiday periods. After forward modelling what would happen in counterfactual scenarios where all or various parts of the vaccination programme were removed, we find that vaccination greatly reduces the influenza-related healthcare burden in England, with an estimated 38,600 to 46,500 hospital admissions averted in 2022/2023 and 55,100 to 64,700 in 2023/2024. Across both seasons, we find that the school part of the programme prevented the most admissions, while in terms of the per-dose impact, the preschool and school parts of the programme were the most effective. Our study highlights vaccine effectiveness (VE) against infection and onwards transmission as a key data gap. In particular, we explore the possibility of lower VE against infection in a sensitivity analysis, and find that the number of averted admissions is reduced to 12,500 to 19,100 in 2022/2023 and 21,900 to 34,700 in 2023/2024.

## 1 Introduction

During the 2022/2023 and 2023/2024 winter seasons, England experienced a significant healthcare burden due to influenza, even with the vaccination programme in place (UK Health Security Agency, 2023, 2024b). For both seasons, the vaccine programme covered everyone aged 65 years and over, individuals at clinical risk older than 6 months of age, preschool and primary school children 2–10 years of age, and secondary school children 11–15 years of age (UK Health Security Agency, 2023, 2024b). For the 2022/2023 season, the vaccine was additionally offered to all 50–64 year olds, which was a temporary programme expansion due to the COVID-19 pandemic, while in secondary schools 11–13 year olds were prioritised depending on vaccine availability (UK Health Security Agency, 2023).

Hospital admissions were high in 2022/2023, reaching the very high Moving Epidemic Method (MEM) threshold (Vega et al., 2013), which was two levels higher than any other season between 2017/2018 and 2023/2024 (UK Health Security Agency, 2024b). It was also the earliest out of these seasons, with hospital admissions breaching the baseline MEM threshold by week 45. However, the season was relatively short, especially when compared to 2017/2018, and there was a significant drop in cases over the Christmas holiday period. This early and high peak is likely a consequence of reduced immunity following the period of low influenza activity during the COVID-19 pandemic (van Leeuwen et al., 2023).

In contrast, 2023/2024 was a more typical season, with hospital admissions breaching the baseline MEM threshold by week 49, before eventually reaching the medium MEM threshold (UK Health Security Agency, 2024b). There were two peaks in the number of cases, which were separated by a reduction over the Christmas holiday period. Both 2022/2023 and 2023/2024 saw relatively low numbers of GP consultations for influenza-like illness (ILI), which could be the result of changes in healthcare seeking behaviour following the COVID-19 pandemic (Read et al., 2023).

In this work, we analyse the impact of the influenza vaccination programme in England on healthcare burden, for the 2022/2023 and 2023/2024 winter seasons. We fit an SEEIIR — Susceptible (S), Exposed (E), Infectious (I) and Recovered (R) — dynamic transmission model to surveillance data, and forward model what would have happened in counterfactual scenarios where various parts of the vaccination programme were removed, including both direct vaccine protection as well as indirect protection of social contacts. Our model is similar to others used previously (Baguelin et al., 2013; van Leeuwen et al., 2017), but with the following updates: (i) in addition to fitting to GP consultation surveillance data, we also fit to hospital admission surveillance data, (ii) we allow for the possibility that vaccine effectiveness (VE) against infection is lower than VE against hospital admission, as seen with COVID-19 (Lopez Bernal et al., 2021; Moore et al., 2021), and (iii) we model changes in the attack rate due to different mixing patterns during school holiday periods.

## 2 Methods

### 2.1 Data

We used Mid-Year Population Estimates for England in 2023 (Office for National Statistics (ONS), 2024), divided into the following age groups: [0, 5), [5, 15), [15, 45), [45, 65) and [65, ∞) (the total population was 57,690,323). Each age group was further divided into ‘low risk’ and ‘high risk’ groups, based on the proportion of individuals at high risk of complications associated with influenza as reported in Immform (Gates et al., 2009). UK contact survey data from the POLYMOD study (Mossong et al., 2008), which contains the number of contacts participants have with different age groups, were accessed using the `socialmixr` R package.

Weekly vaccine uptake stratified by age and risk group was also obtained from Immform. We used estimates for VE against hospital admission shown in Table S1 (Whitaker et al., in preparation), which are based on the methodology described in Whitaker et al. (2024), and assumed that [2, 18) VE applies to the [0, 5) and [5, 15) age groups in our model, and [18, 65) VE to [15, 45) and [45, 65). As was the case in Baguelin et al. (2013), VE against hospital admission is fixed in our model, as opposed to being a fitted parameter in the Bayesian inference. Additionally, the mean is used as a point estimate, since including its uncertainty would come at considerable computational expense (Plummer, 2014).

Data on GP consultations were provided by the Royal College of General Practitioners (RCGP) (UK Health Security Agency, 2023, 2024b), which consists of the weekly number of patients presenting with ILI by age group, as well as the catchment population. In addition, virological sample results were provided for patients that were swabbed. Note that swabs may be taken following ILI or acute respiratory infection diagnosis, but in this analysis we restricted to swabs within two weeks of an ILI diagnosis, as this is more specifically targeted to influenza symptoms.

For hospital admissions, we used Severe Acute Respiratory Infection (SARI) Watch (UK Health Security Agency, 2023, 2024b), which reports the weekly number of laboratory-confirmed influenza hospital admissions in acute National Health Service (NHS) trusts in England, by age group and subtype, as well as the catchment population. The transmission model is fitted to fully subtyped data, however, a significant number of samples in SARI Watch are typed as influenza A but not subtyped further (below we consider these to be subtype ‘AUNK’, i.e., A unknown). To account for this, we assume that the proportion of influenza A that is subtype *s* in each season, *p*_*s*_, is the same as observed in the RCGP virological sample results. Based on this, the underreporting factor for influenza A subtype *s* is given by

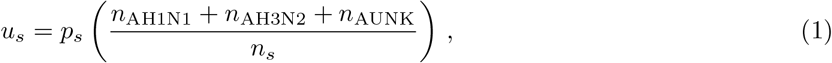

where *n*_*s*_ is the total number of SARI Watch cases reported over a season with subtype *s*. Values for the underreporting factors are listed in Table 1. Note that SARI Watch data has been affected by a step change in influenza case ascertainment following the COVID-19 pandemic (UK Health Security Agency, 2024b), but we do not expect that there has been any significant change over the seasons considered in this analysis.

**Table 1:** Underreporting factors for fully subtyped influenza A hospital admissions in SARI Watch.

| Subtype | 2022/2023 | 2023/2024 |
| --- | --- | --- |
| AH3N2 | 8.88 | 5.49 |
| AH1N1 | 4.38 | 3.81 |

### 2.2 Epidemiological model

Following Baguelin et al. (2013), we model influenza transmission using an age and risk-stratified SEEIIR model, which consists of the set of ordinary differential equations (ODEs) represented schematically in Fig. 1 (For the equations in full, see Eq. (1) in the supplementary information of Baguelin et al., 2013). Here, *S*_*i*_ and 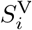 represent the number of non-vaccinated and vaccinated susceptible individuals in cohort *i*, respectively, and analogous for all other compartments, while each cohort *i* corresponds to a particular age group and risk group. The vaccination rate *v*_*i*_ is determined from weekly uptake data (see Sec. 2.1), with a two-week delay added to account for the time taken to develop protective antibodies (Miller et al., 2010). Finally, *α*_*i*_ represents VE against infection, which unlike VE against hospital admission is a fitted parameter in the model (see Sec. 2.3). We assume that there are only three distinct *α*_*i*_ parameters, which are assigned to the five age groups as follows: *α*_1_, *α*_1_, *α*_2_, *α*_2_, *α*_3_.

**Figure 1:**
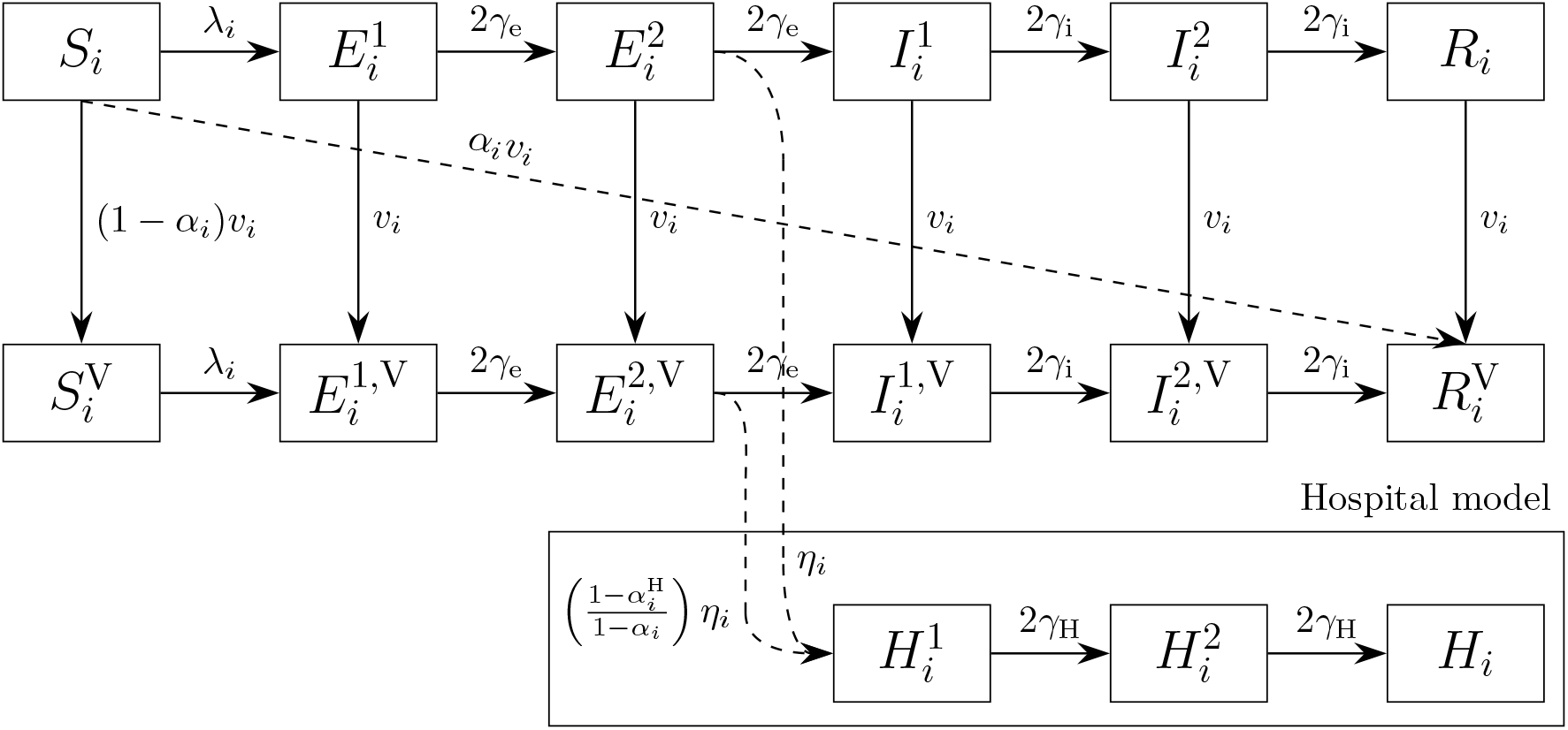
Schematic representation of the compartmental transmission model.

The force of infection is given by

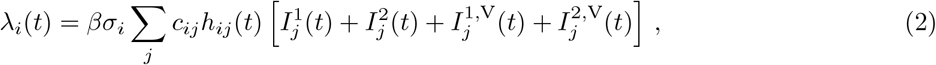

where *β* is the transmission rate, *σ*_*i*_ is the susceptibility, *c*_*ij*_ = *c*_*ji*_ is the contact matrix and *h*_*ij*_(*t*) captures changes in mixing during school holiday periods. Specifically, we assume it takes the form *h*_*ij*_(*t*) = [1 − *h*_*i*_ ℋ(*t*)][1 − *h*_*j*_ℋ(*t*)], where *h*_*i*_ are scaling parameters to be fitted for each cohort *i* and ℋ(*t*) represents the approximate fraction of England where schools are closed each week, based on the `holidayEstR` R package (van Leeuwen & Taylor, 2024). For simplicity, we assume *h*_*i*_ = 0 for all cohorts *i*, except those corresponding to the age groups [5, 15) and [65, ∞). Note that during term time, ℋ(*t*) = 0 hence *h*_*ij*_(*t*) = 1, whereas during school holiday periods positive (negative) *h*_*i*_ indicates lower (higher) mixing.

A new set of ODEs were included in the model to account for hospital admissions (dropping time dependence for clarity):

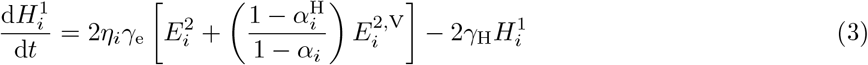

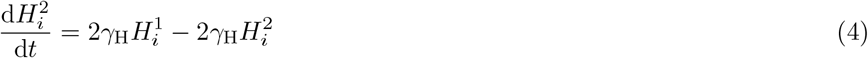

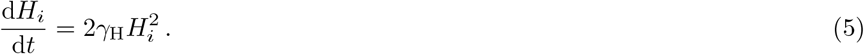

Here, *η*_*i*_ is the infection hospitalisation rate (IHR), 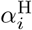 is the VE against hospital admission and *γ*_H_ controls the delay between symptom onset and admission to hospital. The quotient in Eq. (3) reflects the fact that, while the overall vaccine protection against hospital admission is 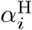, a portion *α*_*i*_ of this was already accounted for in the infection model. Note that we include two delay states 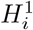 and 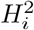, which results in Erlang-2 distributed waiting times between infection and hospitalisation. After fitting the model, in order to estimate the true number of AH1N1 and AH3N2 attributable hospital admissions, the IHR must be scaled up by the underreporting factors of Table 1.

### 2.3 Inference

We fit the epidemiological model to the data using Bayesian inference. Fitting is carried out separately for each season and subtype, using an adaptive Markov Chain Monte Carlo (MCMC) algorithm to generate 2,500 samples from the posterior distribution (ter Braak & Vrugt, 2008).

The likelihood function consists of three parts:

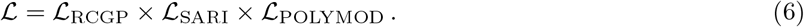

The first part, ℒ_RCGP_, is the GP consultation likelihood based on RCGP data, and is the same as the likelihood function defined by Eq. (10) of Baguelin et al. (2013). The second part is the hospital likelihood, and is given by

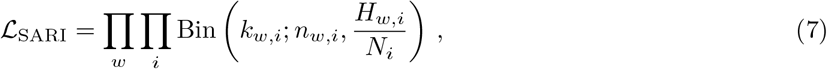

where *k*_*w,i*_ and *n*_*w,i*_ are the number of fully subtyped hospital admissions and the size of the catchment population in week *w* from the SARI Watch data, respectively, *H*_*w,i*_ is the weekly number of new hospital admissions from the transmission model, and *N*_*i*_ represents the total population of England in cohort *i*. Finally, the contact survey likelihood assumes a Poisson distribution and is given by

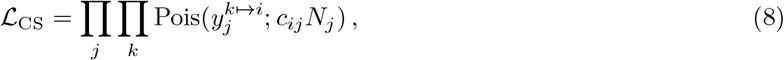

where 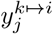 is the number of contacts participant *k*, who belongs to age group *i*, has with age group *j* in the POLYMOD data, and *c*_*ij*_ is the inferred contact matrix, i.e., the rate of a person in age group *i* encountering a person in age group *j*.

We assume flat priors for the contact matrix elements *c*_*ij*_ as well as the school holiday scaling parameters *h*_*i*_. For VE against infection, we use the prior *α*_*i*_ ∼ 1 − LogNormal(−0.61, 0.29) for all seasons, subtypes and age groups, which is obtained by fitting to the odds ratio reported in Dietz et al. (2024) for influenza A/B positivity in those reporting influenza vaccination in both the 2022/2023 and 2021/2022 seasons versus not reporting influenza vaccination in either season. We further require that VE against infection is less than or equal to VE against hospital admission, i.e., 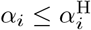, since an individual that is protected from infection is necessarily protected from more severe health outcomes.

To derive the prior for the IHR, *η*_*i*_, we divide samples from uncertainty distributions for the attack rate and number of hospital admissions per person in cohort *i*. For the former, we fit lognormal distributions to seroconversion data from previous seasons (Hayward et al., 2014), whereas the latter are normally distributed based on the mean and 95% confidence interval reported in Table 2 of Cromer et al. (2014). The resulting

**Table 2:** Median number of infections and hospital admissions under the status quo by season and subtype, with 95% confidence interval shown in square brackets.

|  | Subtype | 2022/2023 | 2023/2024 |
| --- | --- | --- | --- |
| Infections (millions) | AH3N2 | 10.3 [9.2 – 11.2] | 7.2 [6.5 – 7.8] |
|  | AH1N1 | 8.4 [7.6 – 9.2] | 9 [8.5 – 9.5] |
|  | B | 4.3 [4 – 4.7] | 3.3 [2.9 – 3.6] |
|  | Total | 23 [21.7 – 24.2] | 19.5 [18.6 – 20.3] |
| Hospital admissions (thousands) | AH3N2 | 28.8 [26 – 31.7] | 15.3 [13.4 – 17.4] |
|  | AH1N1 | 14.3 [11.9 – 16.9] | 25 [22.7 – 27.1] |
|  | B | 7.6 [7.2 – 8] | 3.4 [3.1 – 3.7] |
|  | Total | 50.8 [48.7 – 53.1] | 43.7 [42.2 – 45.1] |

IHR samples are fitted to lognormal distributions, with parameters *µ*_*i*_, *σ*_*i*_. To account for the fact that the transmission model is fitted to fully subtyped data only, the log mean was reduced by the underreporting factors *u*_*s*_ in Table 1 and the log standard deviation was doubled, giving *η*_*i*_ ∼ LogNormal(*µ*_*i*_ − log *u*_*s*_, 2*σ*_*i*_). However, since no seroconversion data were available for the youngest age group, we used a flat prior between 0 and 1%.

### 2.4 Counterfactual scenarios

We define the ‘status quo’ as the scenario where the 2022/2023 and 2023/2024 vaccination programmes are in place, and forward model several counterfactual scenarios where certain cohorts are removed from the status quo programme:

- **R-All**: All individuals are removed from the status quo programme (‘**R-**’ is used to emphasise that the cohort is **removed**)
- **R-Pre**: Individuals below 4 years are removed, i.e., the preschool part of the programme
- **R-Sch**: Individuals aged 4-14 years are removed, i.e., the school part of the programme
- **R-Pae**: Individuals below 15 years are removed, i.e., the paediatric part of the programme (this is a combination of scenarios R-Pre and R-Sch)
- **R-HR**: Individuals aged 15-64 years and at high risk of complications are removed
- **R-50**: Individuals aged 50-64 years and at low risk of complications are removed
- **R-65**: Individuals above 65 years are removed, regardless of risk group

We reran the dynamic transmission model using the same posterior samples as obtained from Bayesian inference, but with the vaccine uptake adjusted to account for the alternative scenarios. To estimate the number of hospital admissions that were prevented by different parts of the vaccination programme, we subtract the number of admissions under the status quo from the number of admissions in each scenario. Since the number of doses distributed under these scenarios differs, we also present the number of hospital admissions averted per additional dose administered.

### 2.5 Main/sensitivity analysis

In the model described so far, VE against infection *α*_*i*_ is an inferred parameter, whose prior distribution is implicitly given equal weight to all other data points. However, this might not be sensible, since the volume of weekly surveillance data means it will have a much larger influence on the fitting than the prior (Spiegelhalter & Best, 2003). To address this, in our main analysis we increase the relative weight of the prior distribution for *α*_*i*_ by a factor of 10, i.e., our main analysis better reflects the available evidence around VE against infection. We also perform a sensitivity analysis that does not include this weight.

## 3 Results

### 3.1 Main analysis

The model fit is shown in Fig. 2 with all age groups combined for simplicity, whereas the full age breakdown is provided in Appendix B. The uncertainty in the data is based on the likelihood function, Eq. (6), i.e., RCGP data follows a hypergeometric distribution (Baguelin et al., 2013) whereas SARI Watch data follows a binomial distribution. Note that influenza A hospital admissions in these figures should not be directly compared with other seasons and subtypes due to underreporting (see Table 1). Including changes in mixing during school holiday periods was particularly impactful for epidemics that started later in the season, namely all types/subtypes in 2023/2024 and influenza B in 2022/2023, with the fits showing a clear double peak structure. Although less obvious, it is still possible to see this effect in the other epidemics, e.g., Fig. S1 shows a small uptick in the [5, 15) age group after schools reopen following the 2022 Christmas holidays.

**Figure 2:**
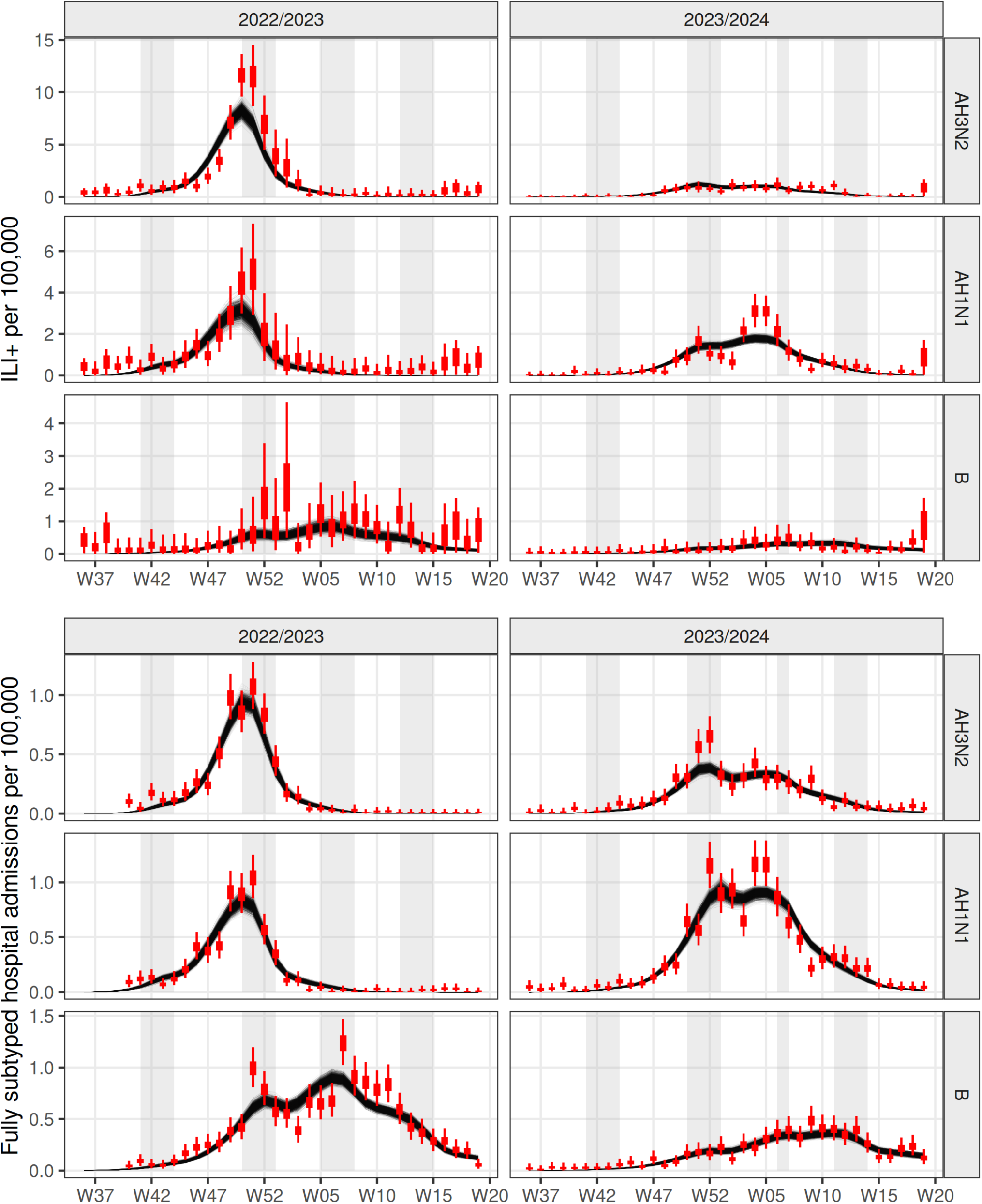
Summary of model fit to weekly GP consultation data (top panel) and weekly hospital admission data (bottom panel) by season and subtype, shown with all age groups combined. In reality, the model is fitted to data by individual age group (see Appendix B). Data is shown in red, black lines represent posterior samples, while the shaded background represents school holiday periods in England.

Indeed, the posterior distributions for *h*_*i*_ are consistent with reduced mixing in the [5, 15) age group during school holiday periods, as shown in Fig. S7. In Fig. 3, we compare the posterior distribution for VE against infection *α*_*i*_ with its prior, and find that the inferred protection is similar to the prior in most cases. While this parameter does not affect the direct impact of the vaccine on hospital admissions, it does affect indirect effects, which will therefore be relatively large (reduced indirect effects are considered in the sensitivity analysis of Sec. 3.2). We also show posterior and prior distributions for the IHR in Fig. S8.

**Figure 3:**
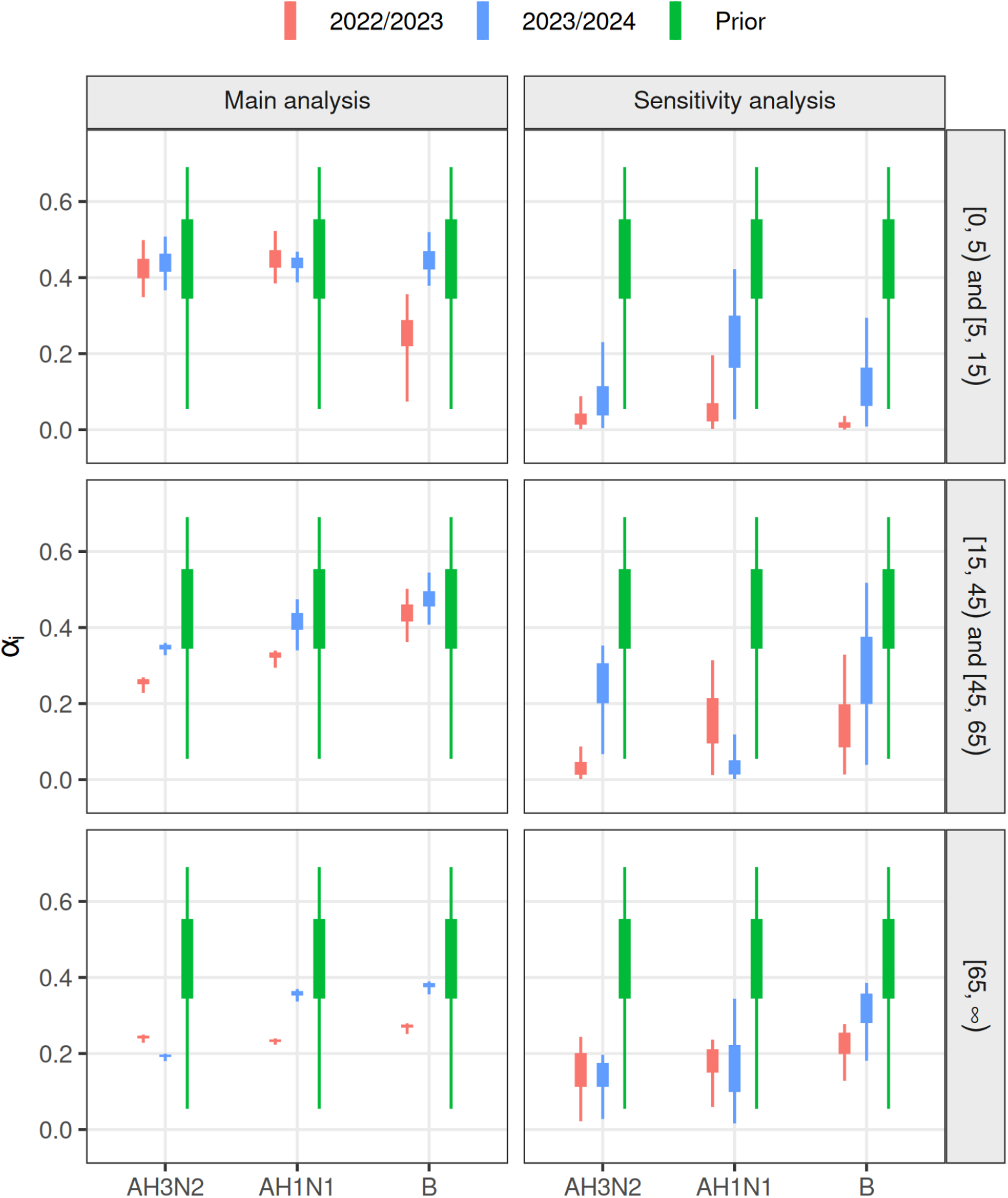
Posterior and prior distributions for VE against infection *α*_*i*_ by season, subtype and age group. In the sensitivity analysis, the prior is given equal weight to all other data points. Plot markers indicate the interquartile range and 95% confidence interval.

The number of infections and hospital admissions under the status quo, based on our fitted model, are shown in Table 2. In 2022/2023, the number of influenza hospital admissions was estimated to range between 48,700 to 53,100, which is consistent with a previous estimate using Secondary Uses Services - Admitted Patient Care (Mellor et al., 2024). The largest burden was due to subtype AH3N2, which contributed just over 50% of the cases, followed by AH1N1 with roughly one third of the cases. Although influenza B had the smallest burden, it was still responsible for a significant number of hospital admissions. There were fewer infections and hospital admissions in 2023/2024, with the latter estimated to range between 42,200 to 45,100. The dominant subtype was instead AH1N1, with over 50% of the cases, whereas roughly one third of the cases were AH3N2 this time.

Fig. 4 shows results for the number of hospital admissions averted under the status quo, compared to when various parts of the vaccination programme are removed (raw numbers are provided in Table S2). Overall, we estimate the number of hospital admissions prevented by the vaccination programme as 38,600 to 46,500 in 2022/2023 and 55,100 to 64,700 in 2023/2024, with the school part of the programme preventing the most admissions in both seasons. We note that the R-Sch and R-50 scenarios changed between 2022/2023 and 2023/2024 (see Sec. 1), which should be taken into account when comparing the two seasons.

**Figure 4:**
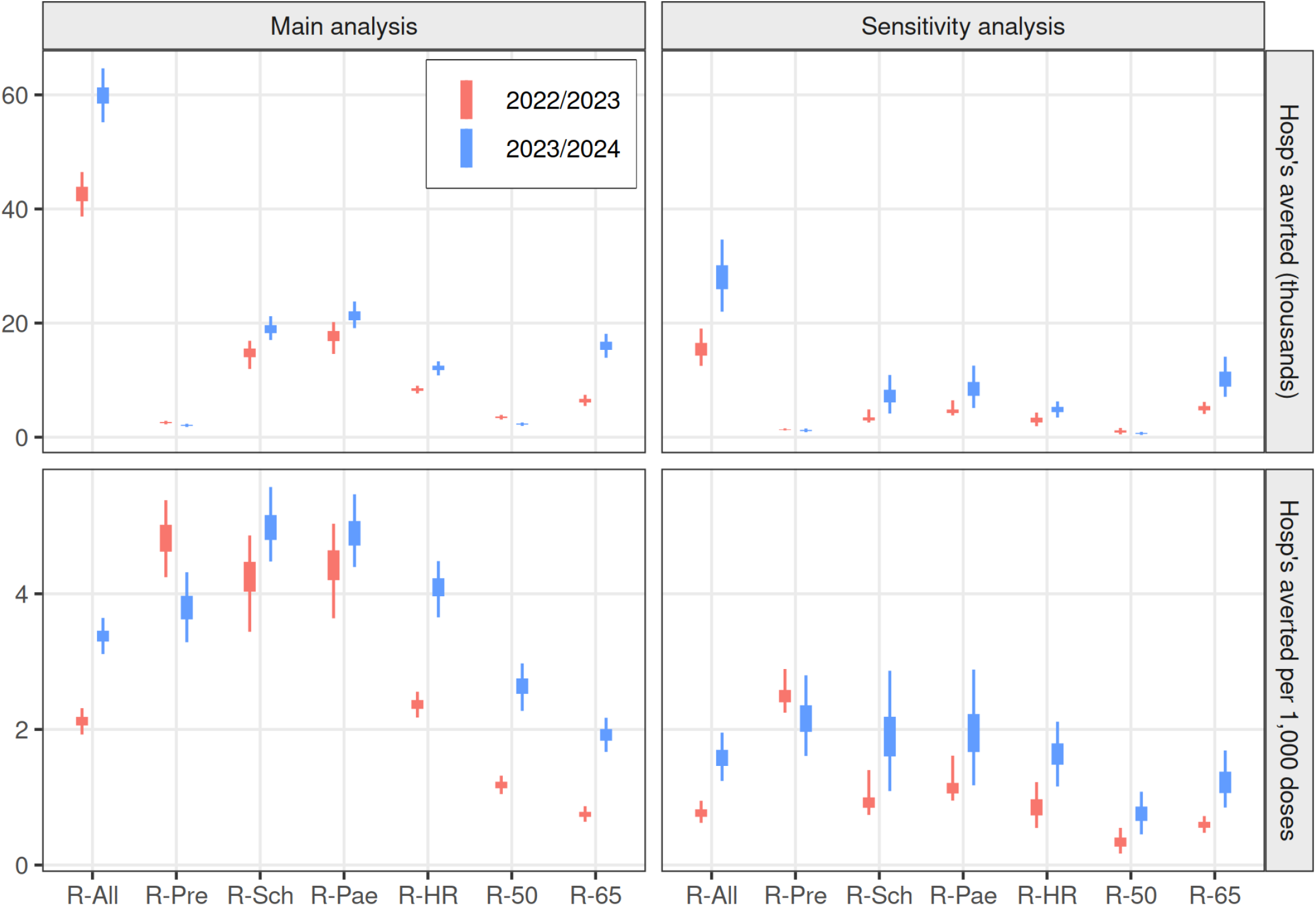
Hospital admissions averted and hospital admissions averted per 1,000 additional doses under the status quo compared to scenarios where various parts of the vaccination programme are removed, for different seasons. In the sensitivity analysis, the prior is given equal weight to all other data points.

Accounting for the number of doses distributed, overall we estimate 1.92 to 2.32 hospital admissions were prevented per 1,000 vaccine doses administered in 2022/2023 and 3.11 to 3.65 in 2023/2024. Most parts of the programme saw an increase in the per-dose impact from 2022/2023 to 2023/2024, which could be explained by improved VE against hospital admission in the [18, 65) and [65, ∞) age groups in relation to the dominant subtype (see Table S1), as well as faster vaccine rollout relative to the timing of the epidemic peak. In both seasons, the preschool and school programmes had some of the highest per-dose impacts. For the former, this is due to the relatively high IHR in the [0, 5) age group (see Fig. S8), whereas the latter is a consequence of the large number of contacts in school-aged children (Mossong et al., 2008), which results in large indirect effects.

### 3.2 Sensitivity analysis

Fig. 3 also shows the posterior distribution for VE against infection when its prior is given equal weight to all other data points. In this case, we find that the inferred protection is significantly lower than the prior in almost all cases, hence indirect effects of vaccination are significantly reduced, in contrast with the main analysis. This has a large effect on the estimated impact of the vaccine programme, as shown in Fig. 4 (raw numbers are provided in Table S3).

Specifically, in the sensitivity analysis we estimate that 12,500 to 19,100 hospital admissions were prevented by the vaccination programme in 2022/2023 and 21,900 to 34,700 in 2023/2024, with the elderly programme preventing the most admissions. In terms of the impact per dose, we find that 0.62 to 0.95 hospital admissions were prevented per 1,000 vaccine doses administered in 2022/2023 and 1.23 to 1.96 in 2023/2024. Per dose, the preschool component is the most impactful part of the programme in the sensitivity analysis, followed by the school component. Hence, even though a large proportion of the benefits of vaccinating school-aged children derive from indirect effects, the direct benefits of vaccination are still significant in this age group.

## 4 Discussion

We fitted a dynamic transmission model to weekly influenza burden data on GP influenza-associated ILI diagnoses and hospital admissions, by season, subtype and age group. The model includes VE against infection and VE against hospital admission as separate parameters, as well as allowing changes in mixing during school holiday periods. Using samples from the posterior distribution, we forward model the healthcare burden assuming different parts of the vaccine programme are omitted. Overall, we find that 38,600 to 46,500 hospital admissions were prevented by the vaccine programme in the 2022/2023 season, and 55,100 to 64,700 in 2023/2024, which is roughly a 50% reduction in cases compared to no programme in both seasons. These are significant numbers, especially in the context of pressures facing the NHS in recent winters (Wise, 2025), and highlight the importance of maintaining high levels of uptake in the face of persistent decline in certain target groups, e.g., pregnant women and healthcare workers (UK Health Security Agency, 2024a).

Two previous studies have used a similar model to explore the impact of the influenza vaccination programme. Under an elderly and high risk programme, 6,800 to 19,200 (Baguelin et al., 2015) and 9,800 to 20,700 (Wenzel et al., 2021) yearly hospital admissions were predicted. Further adding a paediatric programme covering all 2 to 16 year olds reduced the burden to 2,500 to 10,500 (Baguelin et al., 2015) and 3,500 to 7,500 (Wenzel et al., 2021), which translated into a reduction in mean yearly hospital admissions of 6,100 and 8,700, respectively. In comparison, we estimate an increase of hospital admissions in the absence of the current paediatric programme between 14,500 to 20,200 in 2022/2023 and 19,100 to 23,800 in 2023/2024 (in the sensitivity analysis, 3,800 to 6,500 in 2022/2023 and 5,100 to 12,600 in 2023/2024). Direct comparison of these numbers is difficult due to differences in the models. For example, previous work averaged over a large number of seasons, whereas we have only considered two seasons with relatively high burden. Moreover, the paediatric programme was different in 2022/2023, not covering the two oldest school years. The models also make different assumptions around vaccine protection against infection.

The model results in a high influenza attack rate, especially in 2022/2023, which is mainly driven by our estimation of the IHR using historic seroconversion and influenza-related hospital admissions data (see Appendix 2.3). While the attack rate of all three subtypes together was higher than historically found (Hayward et al., 2014), it is not outside the range of possibilities (Cohen et al., 2021), especially given the lack of influenza during the COVID-19 pandemic and the potential impact on population immunity. However, in most cases we found that the IHR posteriors were higher than the priors (see Fig. S8), which would suggest that our priors are too low and contribute to an overestimation of the attack rate. If our attack rate is too high, we are likely underestimating the indirect impact of vaccination on hospital admissions, because when prevalence is high individuals will encounter many unprotected individuals and one is likely to be infected. However, direct impact will scale linearly with the number of hospital admissions in a given year, so is robust to changes in the attack rate.

As highlighted by this study, an important open question is the protection of influenza vaccines versus infection and onward transmission, as compared to more severe clinical endpoints such as hospital admission. Previous work did not consider these as separate model parameters (Backer et al., 2018; Baguelin et al., 2013; Hill et al., 2019; Pitman et al., 2013; Sandmann et al., 2022; Wenzel et al., 2021), since there were no data available on protection against infection only, but this is unlikely to be a good assumption based on the COVID-19 pandemic (Lopez Bernal et al., 2021; Moore et al., 2021). Here, our sensitivity analysis used lower estimates of VE against infection, and this showed that estimates of the impact of the vaccination programme are very sensitive to this assumption. This was particularly true for parts of the programme that target individuals with more contacts, e.g., school-aged children, although we note that even under low protection against infection we still found that the paediatric programme had a significant impact on hospital admissions.

There are several further areas for development of this model. Firstly, we have only considered hospital admissions prevented by the vaccination programme, but averted mortality is another benefit that could be estimated. This would be necessary for a cost-effectiveness analysis, where mortality makes a significant contribution to the overall burden in terms of Quality-Adjusted Life Years (Baguelin et al., 2015). Second, the POLYMOD contact survey data is almost two decades old (Mossong et al., 2008), and it is possible that mixing patterns have changed during this time, especially following the COVID-19 pandemic. A new UK social contact survey is currently ongoing, which could be used in future versions of this model. Related to this point, contact parameters in the model (*c*_*ij*_, *h*_*i*_) are fitted separately for each subtype (see Fig. S7), even though contact patterns are subtype independent. Hence, an alternative approach would be to fit all three subtypes simultaneously using a common set of contact parameters, significantly reducing the parameter space of the model. Finally, our model outputs do not include uncertainty in VE against hospital admission, and we do not account for any waning of immunity following vaccination, even though data does exist that could allow both of these things (Whitaker et al., in preparation).

## Data Availability

All data produced in the present study are available upon reasonable request to the authors

## Acknowledgments

The authors would like to thank Heather Whitaker and Freja Kirsebom for contributing VE data, Suzanne Elgohari for providing SARI Watch data, Catherine Quinot for providing RCGP data, and Katja Hoschler and Jamie Lopez Bernal for helpful discussions.

The views expressed are those of the authors and not necessarily those of DHSC or UKHSA. UKHSA undertakes postmarketing surveillance and regulatory analyses for vaccine makers for which it charges cost recovery payments.

## A VE against hospital admission

**Table S1:**
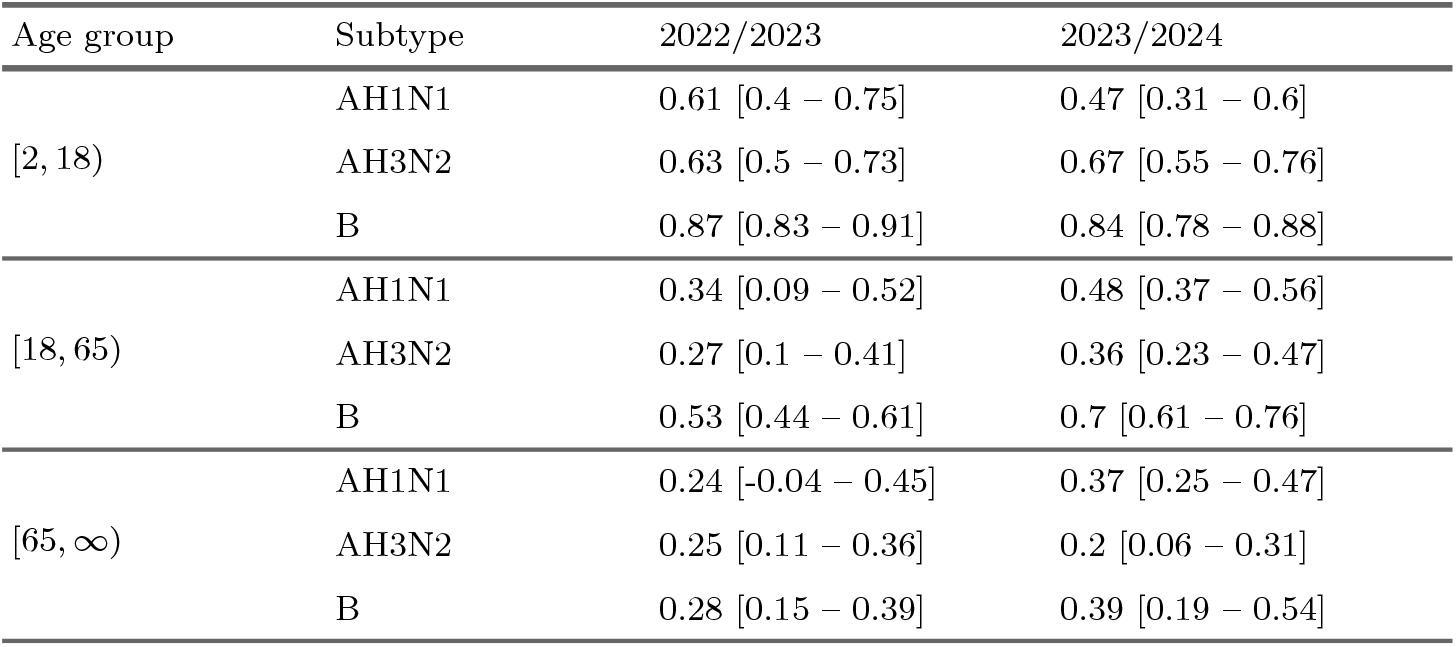
Mean VE against hospital admission by season, sub-type and age group, with 95% confidence interval shown in square brackets. Taken from (Whitaker et al., in preparation).

## B Inference results

**Figure S1:**
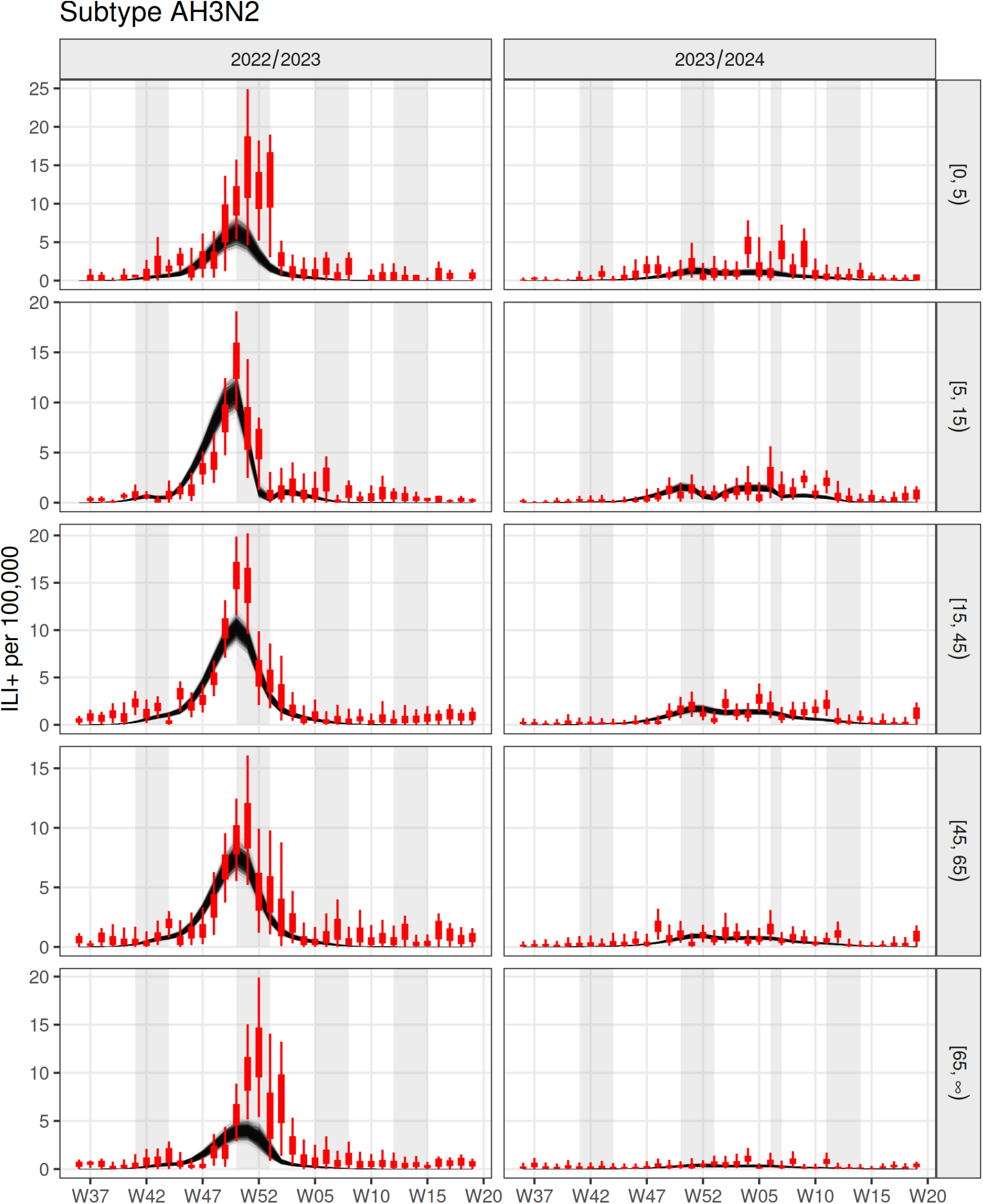
Model fit to weekly GP consultation data by season, for subtype AH3N2. Data is shown in red, black lines represent posterior samples, while the shaded background represents school holiday periods in England.

**Figure S2:**
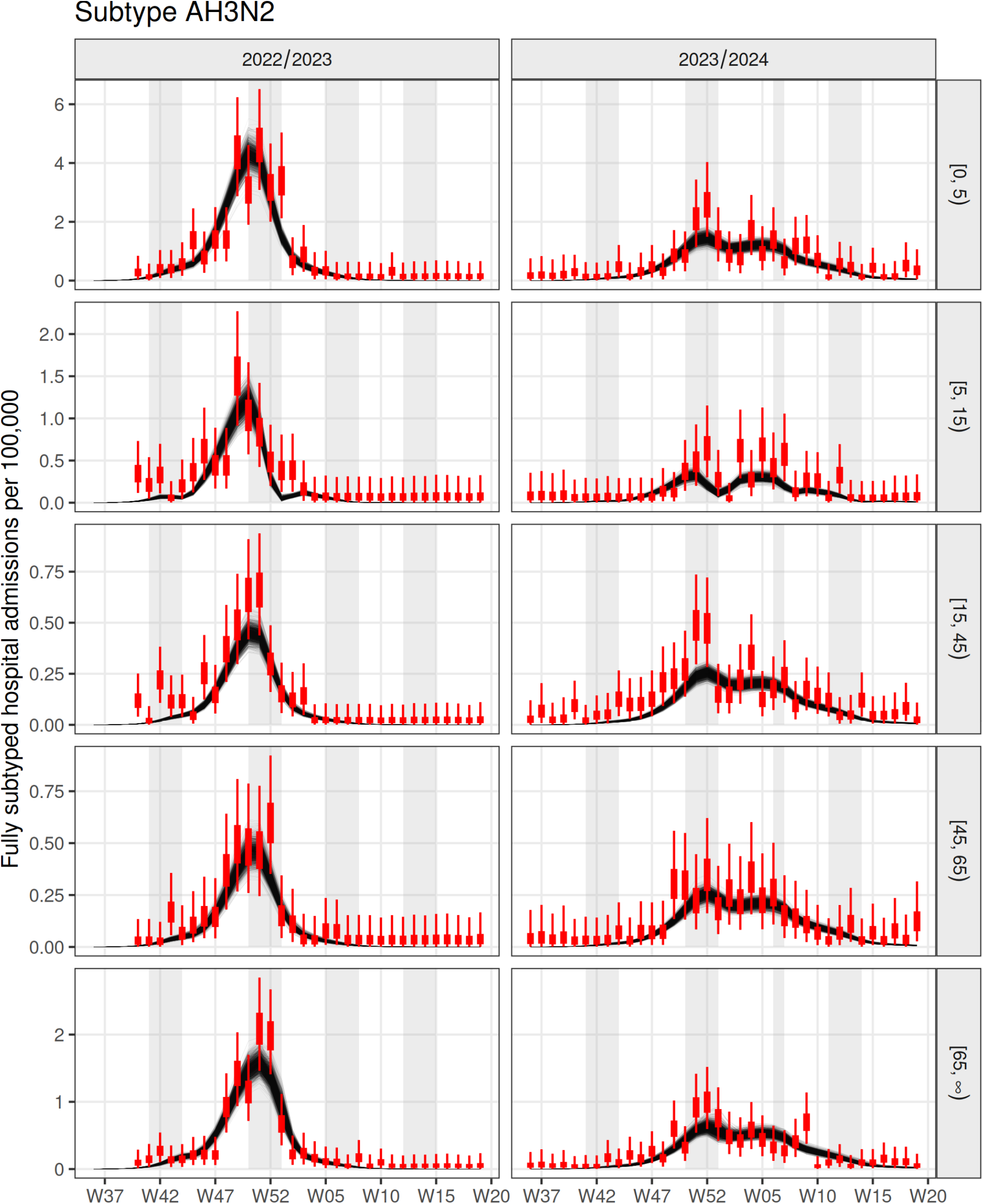
Model fit to weekly hospital admission data by season, for subtype AH3N2. Data is shown in red, black lines represent posterior samples, while the shaded background represents school holiday periods in England.

**Figure S3:**
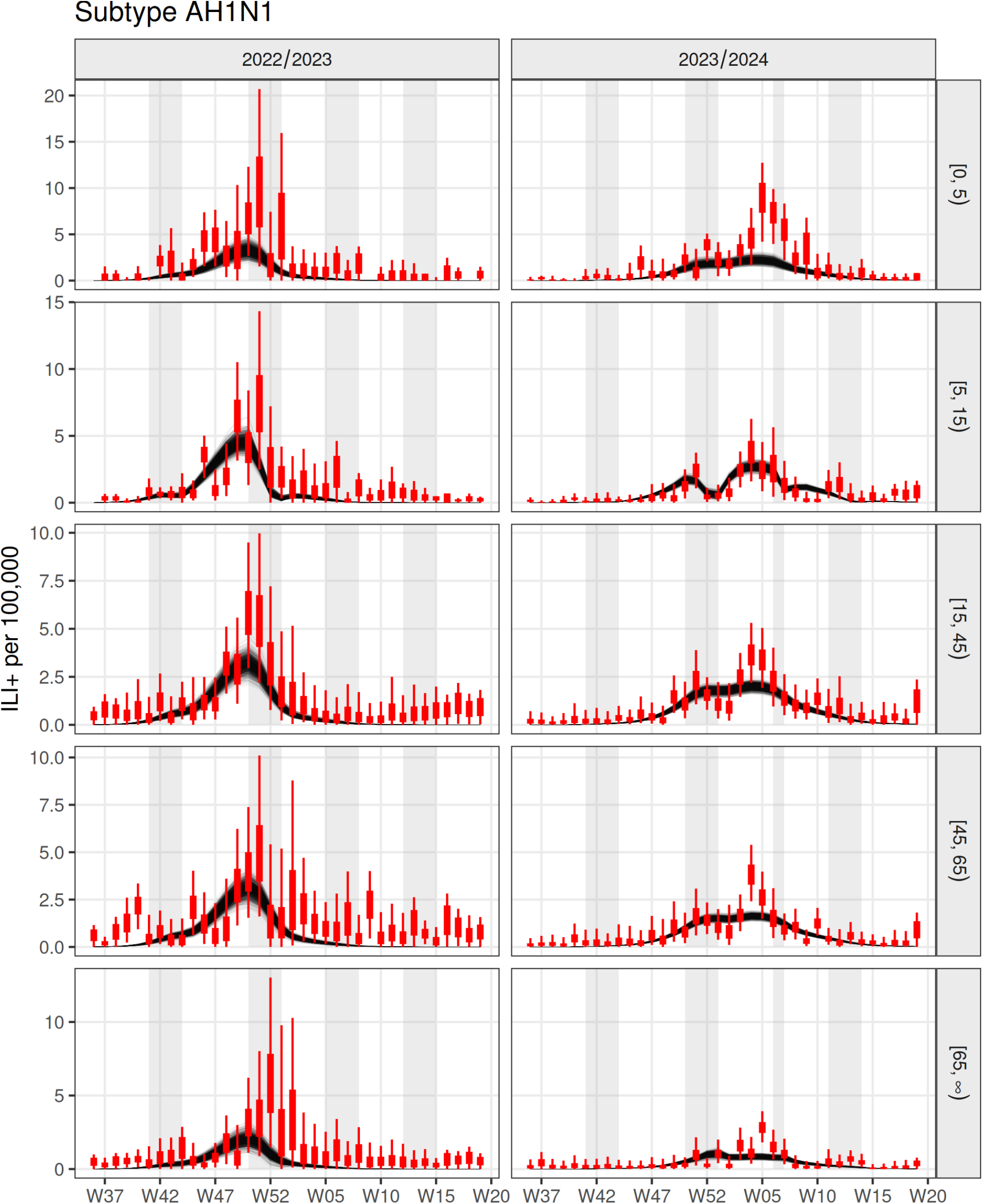
Model fit to weekly GP consultation data by season, for subtype AH1N1. Data is shown in red, black lines represent posterior samples, while the shaded background represents school holiday periods in England.

**Figure S4:**
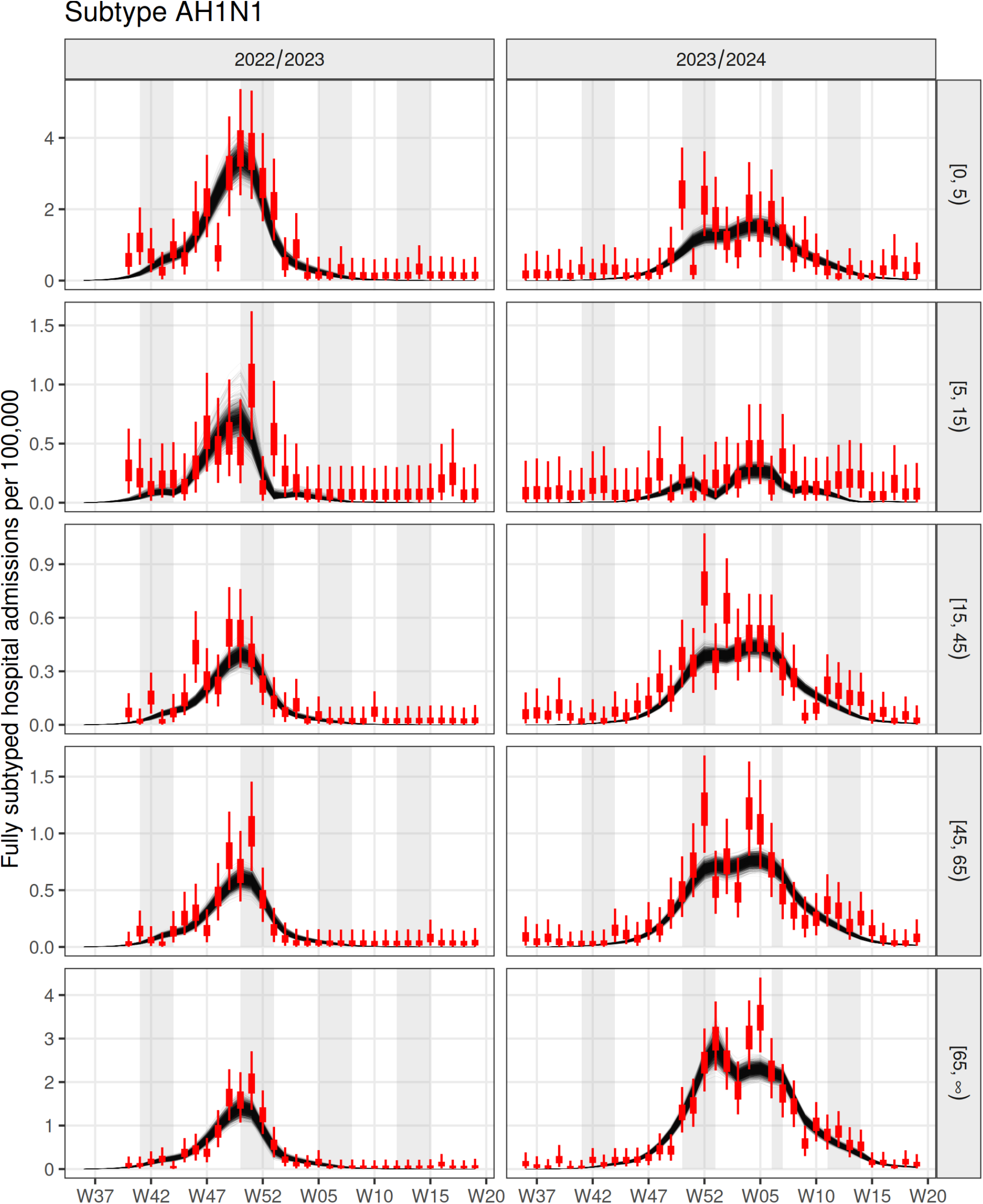
Model fit to weekly hospital admission data by season, for subtype AH1N1. Data is shown in red, black lines represent posterior samples, while the shaded background represents school holiday periods in England.

**Figure S5:**
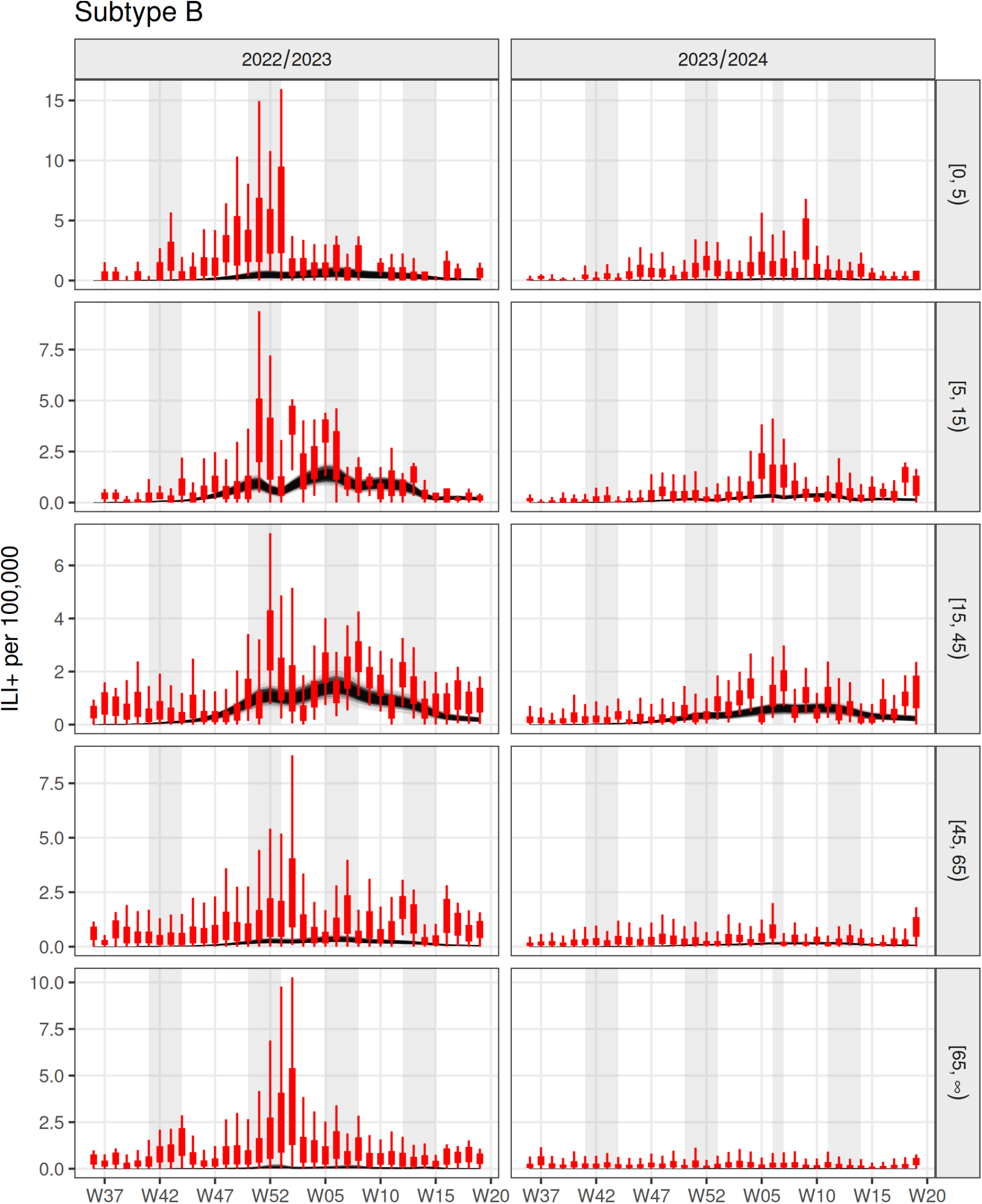
Model fit to weekly GP consultation data by season, for subtype B. Data is shown in red, black lines represent posterior samples, while the shaded background represents school holiday periods in England.

**Figure S6:**
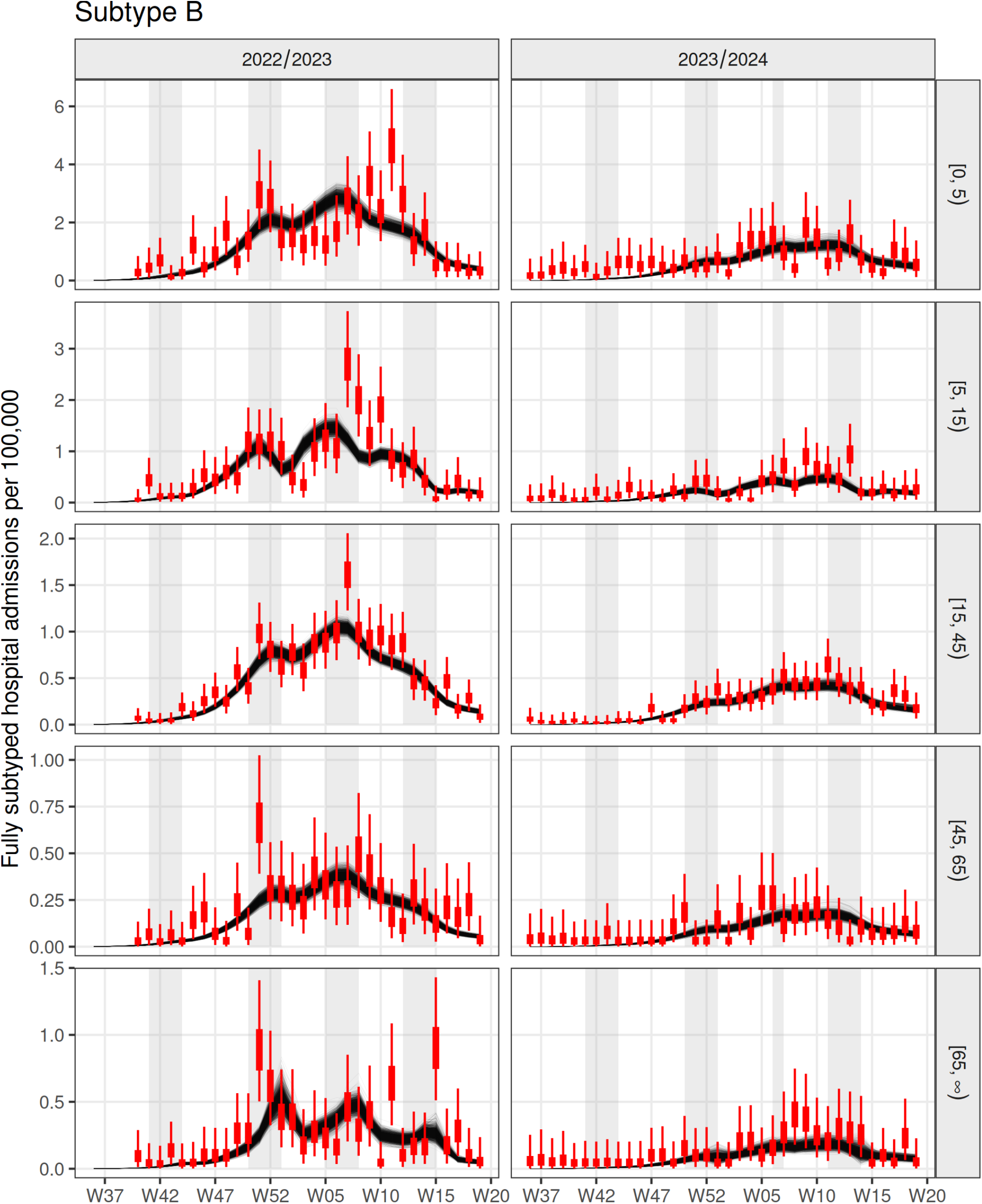
Model fit to weekly hospital admission data by season, for subtype B. Data is shown in red, black lines represent posterior samples, while the shaded background represents school holiday periods in England.

**Figure S7:**
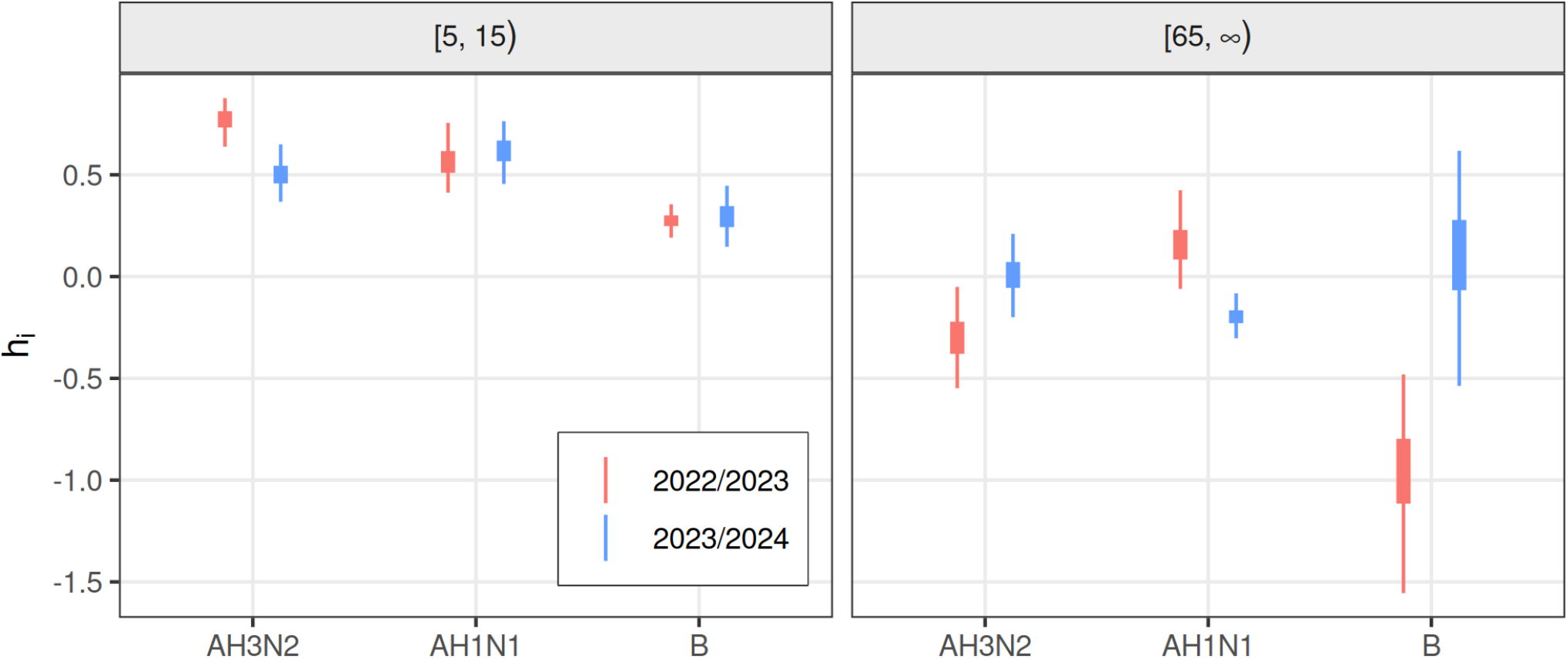
Posterior distributions for the school holiday scaling parameters *h*_*i*_ by season, subtype and age group.

**Figure S8:**
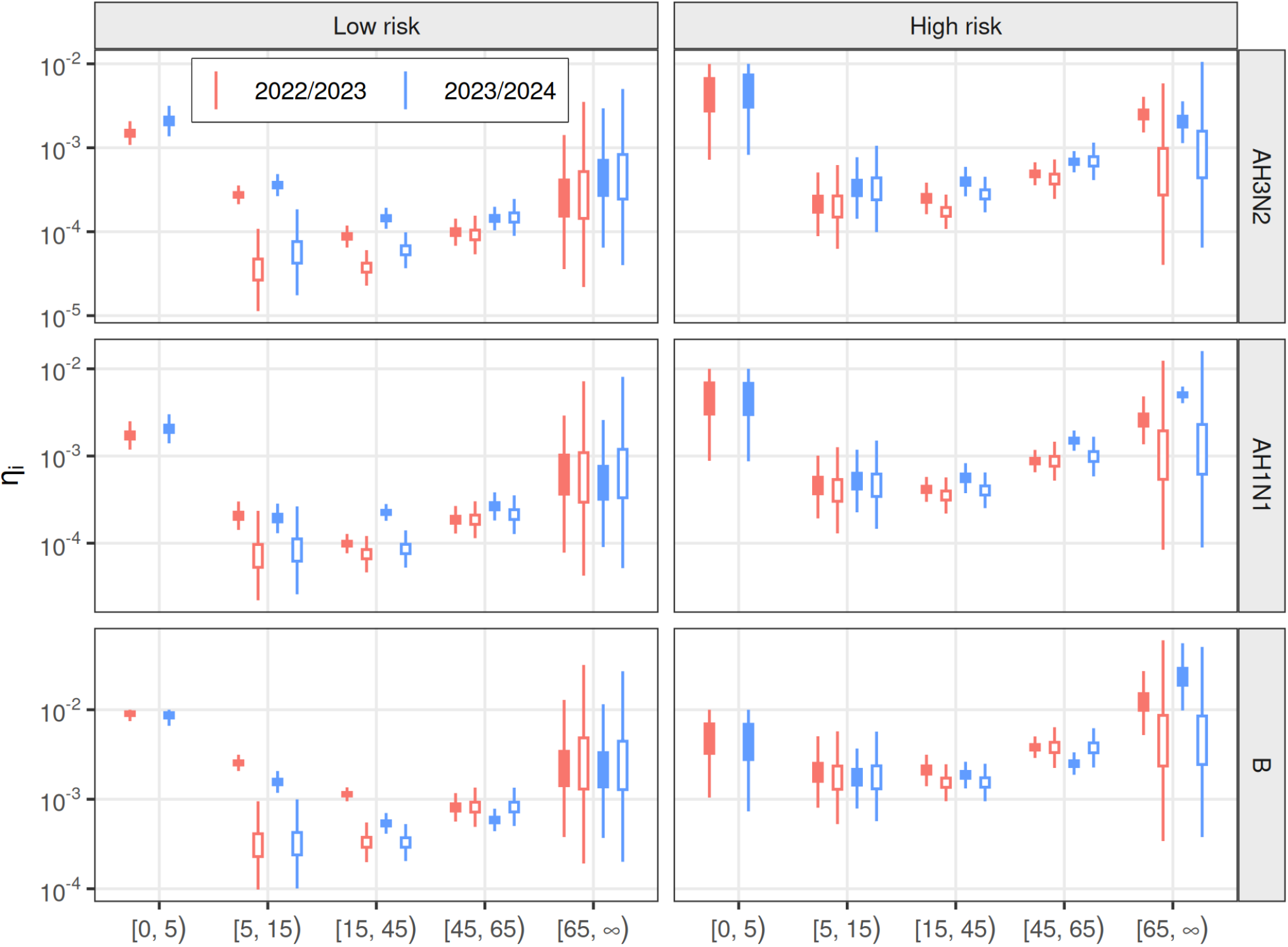
Posterior and prior distributions for the IHR *η*_*i*_ by season, subtype, age group and risk group. Filled and hollow plot markers represent posterior and prior distributions, respectively.

## C Hospital admissions averted

**Table S2:** Main analysis.

|  | Scenario | 2022/2023 | 2023/2024 |
| --- | --- | --- | --- |
| Hospital admissions averted<br>(thousands) | R-All | 42.6 [38.6 – 46.5] | 59.9 [55.1 – 64.7] |
|  | R-Pre | 2.6 [2.2 – 2.9] | 2.1 [1.7 – 2.4] |
|  | R-Sch | 14.8 [11.9 – 16.9] | 18.9 [17 – 21.3] |
|  | R-Pae | 17.8 [14.5 – 20.2] | 21.2 [19.1 – 23.8] |
|  | R-HR | 8.4 [7.6 – 9.1] | 12.1 [10.8 – 13.3] |
|  | R-50 | 3.5 [3 – 3.9] | 2.3 [1.9 – 2.6] |
|  | R-65 | 6.4 [5.4 – 7.5] | 16 [13.9 – 18.2] |
| Hospital admissions averted<br>per 1,000 doses | R-All | 2.12 [1.92 – 2.32] | 3.38 [3.11 – 3.65] |
|  | R-Pre | 4.82 [4.24 – 5.39] | 3.8 [3.28 – 4.32] |
|  | R-Sch | 4.26 [3.43 – 4.87] | 4.97 [4.47 – 5.58] |
|  | R-Pae | 4.44 [3.63 – 5.04] | 4.88 [4.39 – 5.47] |
|  | R-HR | 2.37 [2.17 – 2.56] | 4.09 [3.65 – 4.49] |
|  | R-50 | 1.18 [1.04 – 1.32] | 2.64 [2.27 – 2.98] |
|  | R-65 | 0.75 [0.63 – 0.87] | 1.92 [1.66 – 2.18] |

**Table S3:** Sensitivity analysis.

|  | Scenario | 2022/2023 | 2023/2024 |
| --- | --- | --- | --- |
| Hospital admissions averted<br>(thousands) | R-All | 15.4 [12.5 – 19.1] | 28 [21.9 – 34.7] |
|  | R-Pre | 1.3 [1.1 – 1.6] | 1.2 [0.8 – 1.6] |
|  | R-Sch | 3.2 [2.5 – 4.9] | 7.2 [4.1 – 11] |
|  | R-Pae | 4.5 [3.8 – 6.5] | 8.5 [5.1 – 12.6] |
|  | R-HR | 3 [1.9 – 4.4] | 4.9 [3.4 – 6.3] |
|  | R-50 | 1 [0.4 – 1.7] | 0.7 [0.3 – 1] |
|  | R-65 | 5.1 [4 – 6.2] | 10 [7 – 14.2] |
| Hospital admissions averted<br>per 1,000 doses | R-All | 0.76 [0.62 – 0.95] | 1.58 [1.23 – 1.96] |
|  | R-Pre | 2.48 [2.24 – 2.9] | 2.15 [1.6 – 2.8] |
|  | R-Sch | 0.91 [0.73 – 1.41] | 1.9 [1.09 – 2.87] |
|  | R-Pae | 1.13 [0.94 – 1.62] | 1.95 [1.17 – 2.89] |
|  | R-HR | 0.85 [0.54 – 1.23] | 1.64 [1.15 – 2.12] |
|  | R-50 | 0.33 [0.16 – 0.55] | 0.76 [0.45 – 1.09] |
|  | R-65 | 0.59 [0.47 – 0.73] | 1.2 [0.84 – 1.7] |

## References

Backer, J. A., Wallinga, J., Meijer, A., Donker, G. A., van der Hoek, W., & van Boven, M. (2018). The impact of influenza vaccination on infection, hospitalisation and mortality in the Netherlands between 2003 and 2015. Epidemics. https://doi.org/ggvp9m

Baguelin, M., Camacho, A., Flasche, S., & Edmunds, J. W. (2015). Extending the elderly- and risk-group programme of vaccination against seasonal influenza in England and Wales: A cost-effectiveness study. BMC Medicine, 13 (1). https://doi.org/f7t324

Baguelin, M., Flasche, S., Camacho, A., Demiris, N., Miller, E., & Edmunds, W. J. (2013). Assessing Optimal Target Populations for Influenza Vaccination Programmes: An Evidence Synthesis and Modelling Study. PLoS Med, 10 (10), e1001527. https://doi.org/gbfntv

Cohen, C., Kleynhans, J., Moyes, J., McMorrow, M. L., Treurnicht, F. K., Hellferscee, O., Mathunjwa, A., Gottberg, A. von, Wolter, N., Martinson, N. A., Kahn, K., Lebina, L., Mothlaoleng, K., Wafawanaka, F., Gómez-Olivé, F. X., Mkhencele, T., Mathee, A., Piketh, S., Language, B., … PHIRST group. (2021). Asymptomatic transmission and high community burden of seasonal influenza in an urban and a rural community in South Africa, 2017-18 (PHIRST): A population cohort study. Lancet Glob. Health, 9 (6), e863–e874.

Cromer, D., van Hoek, A. J., Jit, M., Edmunds, W. J., Fleming, D., & Miller, E. (2014). The burden of influenza in England by age and clinical risk group: A statistical analysis to inform vaccine policy. The Journal of Infection, 68 (4), 363–371. https://doi.org/f2qh5d

Dietz, E., Pritchard, E., Pouwels, K., Ehsaan, M., Blake, J., Gaughan, C., Haduli, E., Boothe, H., Vihta, K.-D., Peto, T., Stoesser, N., Matthews, P., Taylor, N., Diamond, I., Studley, R., Rourke, E., Birrell, P., De Angelis, D., Fowler, T., … Walker, A. S. (2024). SARS-CoV-2, influenza A/B and respiratory syncytial virus positivity and association with influenza-like illness and self-reported symptoms, over the 2022/23 winter season in the UK: A longitudinal surveillance cohort. BMC Medicine, 22 (1). 10.1186/s12916-024-03351-w

Gates, P., Noakes, K., Begum, F., Pebody, R., & Salisbury, D. (2009). Collection of routine national seasonal influenza vaccine coverage data from GP practices in England using a web-based collection system. Vaccine, 27 (48), 6669–6677. 10.1016/j.vaccine.2009.08.094

Hayward, A. C., Fragaszy, E. B., Bermingham, A., Wang, L., Copas, A., Edmunds, W. J., Ferguson, N., Goonetilleke, N., Harvey, G., Kovar, J., Lim, M. S. C., McMichael, A., Millett, E. R. C., Nguyen-Van-Tam, J. S., Nazareth, I., Pebody, R., Tabassum, F., Watson, J. M., Wurie, F. B., … Flu Watch Group. (2014). Comparative community burden and severity of seasonal and pandemic influenza: Results of the Flu Watch cohort study. The Lancet. Respiratory Medicine, 2 (6), 445–454. https://doi.org/f3hxg5

Hill, E. M., Petrou, S., Lusignan, S. de, Yonova, I., & Keeling, M. J. (2019). Seasonal influenza: Modelling approaches to capture immunity propagation. PLOS Computational Biology, 15 (10), e1007096. https://doi.org/ghfqrm

Lopez Bernal, J., Andrews, N., Gower, C., Robertson, C., Stowe, J., Tessier, E., Simmons, R., Cottrell, S., Roberts, R., O’Doherty, M., Brown, K., Cameron, C., Stockton, D., McMenamin, J., & Ramsay, M. (2021). Effectiveness of the Pfizer-BioNTech and Oxford-AstraZeneca vaccines on covid-19 related symptoms, hospital admissions, and mortality in older adults in England: Test negative case-control study. BMJ, 1088. 10.1136/bmj.n1088

Mellor, J., Christie, R., Guilder, J., Paton, R. S., Elgohari, S., Watson, C., Deeny, S. R., & Ward, T. (2024). A comparative study of influenza surveillance systems and administrative data in England during the 2022–2023 season. PLOS Global Public Health, 4 (9), e0003627. 10.1371/journal.pgph.000362

Miller, E., Hoschler, K., Hardelid, P., Stanford, E., Andrews, N., & Zambon, M. (2010). Incidence of 2009 pandemic influenza A H1N1 infection in England: A cross-sectional serological study. The Lancet, 375 (9720), 1100–1108. 10.1016/S0140-6736(09)62126-7

Moore, S., Hill, E. M., Tildesley, M. J., Dyson, L., & Keeling, M. J. (2021). Vaccination and non-pharmaceutical interventions for COVID-19: A mathematical modelling study. The Lancet Infectious Diseases, 21 (6), 793–802. 10.1016/s1473-3099(21)00143-2

Mossong, J., Hens, N., Jit, M., Beutels, P., Auranen, K., Mikolajczyk, R., Massari, M., Salmaso, S., Tomba, G., Wallinga, J., Heijne, J., Sadkowska-Todys, M., Rosinska, M., & Edmunds, W. J. (2008). Social contacts and mixing patterns relevant to the spread of infectious diseases. PLoS Medicine, 5 (3), e74. https://doi.org/ch2stb

Office for National Statistics (ONS). (2024). Estimates of the population for the UK, England, Wales, Scotland, and Northern Ireland. https://www.ons.gov.uk/peoplepopulationandcommunity/populationandmigration/populationestimates/datasets/populationestimatesforukenglandandwalesscotlandandnorthernireland.

Pitman, R. J., Nagy, L. D., & Sculpher, M. J. (2013). Cost-effectiveness of childhood influenza vaccination in England and Wales: Results from a dynamic transmission model. Vaccine, 31 (6), 927–942. https://doi.org/f2ns8f

Plummer, M. (2014). Cuts in Bayesian graphical models. Statistics and Computing, 25 (1), 37–43. 10.1007/s11222-014-9503-z

Read, B., McLeod, M., Tonkin-Crine, S., Ashiru-Oredope, D., Quigley, A., Brown, C. S., & Lecky, D. M. (2023). Changes in public health-seeking behaviours for self-limiting respiratory tract infections across england during the COVID-19 pandemic. European Journal of Public Health, 33 (6), 987–993. 10.1093/eurpub/ckad136

Sandmann, F. G., van Leeuwen, E., Bernard-Stoecklin, S., Casado, I., Castilla, J., Domegan, L., Gherasim, A., Hooiveld, M., Kislaya, I., Larrauri, A., Levy-Bruhl, D., Machado, A., Marques, D. F. P., Martínez-Baz, I., Mazagatos, C., McMenamin, J., Meijer, A., Murray, J. L. K., Nunes, B., … Baguelin, M. (2022). Health and economic impact of seasonal influenza mass vaccination strategies in European settings: A mathematical modelling and cost-effectiveness analysis. Vaccine, 40 (9), 1306–1315. 10.1016/j.vaccine.2022.01.015

Spiegelhalter, D. J., & Best, N. G. (2003). Bayesian approaches to multiple sources of evidence and uncertainty in complex cost-effectiveness modelling. Statistics in Medicine, 22 (23), 3687–3709. 10.1002/sim.1586

ter Braak, C. J. F., & Vrugt, J. A. (2008). Differential Evolution Markov Chain with snooker updater and fewer chains. Statistics and Computing, 18 (4), 435–446. https://doi.org/dcdqpb

UK Health Security Agency. (2023). Surveillance of influenza and other seasonal respiratory viruses in the UK, winter 2022 to 2023. gov.uk. https://www.gov.uk/government/statistics/annual-flu-reports/surveillance-of-influenza-and-other-seasonal-respiratory-viruses-in-the-uk-winter-2022-to-2023

UK Health Security Agency. (2024a). Seasonal influenza vaccine uptake in GP patients in England: Winter season 2023 to 2024. gov.uk. https://www.gov.uk/government/statistics/seasonal-influenza-vaccine-uptake-in-gp-patients-winter-season-2023-to-2024/seasonal-influenza-vaccine-uptake-in-gp-patients-in-england-winter-season-2023-to-2024

UK Health Security Agency. (2024b). Surveillance of influenza and other seasonal respiratory viruses in the UK, winter 2023 to 2024. gov.uk. https://www.gov.uk/government/statistics/surveillance-of-influenza-and-other-seasonal-respiratory-viruses-in-the-uk-winter-2023-to-2024/surveillance-of-influenza-and-other-seasonal-respiratory-viruses-in-the-uk-winter-2023-to-2024

van Leeuwen, E., Klepac, P., Thorrington, D., Pebody, R. G., & Baguelin, M. (2017). fluEvidenceSynthesis: An R package for evidence synthesis based analysis of epidemiological outbreaks. PLOS Computational Biology, 13 (11), e1005838. https://doi.org/gck6wz

van Leeuwen, E., Panovska-Griffiths, J., Elgohari, S., Charlett, A., & Watson, C. (2023). The interplay between susceptibility and vaccine effectiveness control the timing and size of an emerging seasonal influenza wave in England. Epidemics, 44, 100709. 10.1016/j.epidem.2023.100709

van Leeuwen, E., & Taylor, T. (2024). holidayEstR. In GitLab repository. https://gitlab.com/epidemics-r/holidayestr; GitLab.

Vega, T., Lozano, J. E., Meerhoff, T., Snacken, R., Mott, J., Lejarazu, R. O. de, & Nunes, B. (2013). Influenza surveillance in Europe: Establishing epidemic thresholds by the Moving Epidemic Method. Influenza and Other Respiratory Viruses, 7 (4), 546–558. https://doi.org/ggvp9s

Wenzel, N. S., Atkins, K. E., van Leeuwen, E., Halloran, M. E., & Baguelin, M. (2021). Cost-effectiveness of live-attenuated influenza vaccination among school-age children. Vaccine, 39 (2), 447–456. https://doi.org/gj6bn3

Whitaker, H. J., Hassell, K., Hoschler, K., Power, L., Stowe, J., Boddington, N. L., Tsang, C., Zhao, H., Linley, E., Button, E., Okusi, C., Aspden, C., Byford, R., deLusignan, S., Amirthalingam, G., Zambon, M., Andrews, N. J., & Watson, C. (2024). Influenza vaccination during the 2021/22 season: A data-linkage test-negative case-control study of effectiveness against influenza requiring emergency care in England and serological analysis of primary care patients. Vaccine, 42 (7), 1656–1664. 10.1016/j.vaccine.2024.02.006

Whitaker, H. J., Kirsebom, F. C. M., Hassell, K., Quinot, C., Stowe, J., Hoschler, K., Zambon, M., Andrews, N. J., Watson, C. H., & Lopez Bernal, J. (in preparation). Relative effectiveness of influenza vaccines and duration of protection against hospitalisation in England: 2022/23 and 2023/24 seasons.

Wise, J. (2025). Winter pressure: NHS struggles to cope with flu surge as hospitals declare critical incidents. BMJ, 388. 10.1136/bmj.r51

